# Forecasting the dynamics of COVID-19 Pandemic in Top 15 countries in April 2020: ARIMA Model with Machine Learning Approach

**DOI:** 10.1101/2020.03.30.20046227

**Authors:** Pavan Kumar, Himangshu Kalita, Shashikanta Patairiya, Yagya Datt Sharma, Chintan Nanda, Meenu Rani, Jamal Rahmani, Akshaya Srikanth Bhagavathula

## Abstract

We here predicted some trajectories of COVID-19 in the coming days (until April 30, 2020) using the most advanced Auto-Regressive Integrated Moving Average Model (ARIMA). Our analysis predicted very frightening outcomes, which defines to worsen the conditions in Iran, entire Europe, especially Italy, Spain, and France. While South Korea, after the initial blast, has come to stability, the same goes for the COVID-19 origin country China with more positive recovery cases and confirm to remain stable. The United States of America (USA) will come as a surprise and going to become the epicenter for new cases during the mid-April 2020. Based on our predictions, public health officials should tailor aggressive interventions to grasp the power exponential growth, and rapid infection control measures at hospital levels are urgently needed to curtail the COVID-19 pandemic.

---

Since January 2020, the novel coronavirus (severe acute respiratory syndrome 2 [SARS-CoV-2]) from Wuhan, China, has continued to spread around the world and turned into Pandemic as novel coronavirus disease (COVID-19) [1-2]. Due to rapid pandemic potential and the absence of vaccines and drugs, the contagious COVID-19 disease devastated the normal life across the globe. Currently, Coronavirus infected more than half a million population, and killed more than 25 thousand people, and also forced more than 3 billion to stay with their homes [3]. Due to a lack of knowledge about this virus, the COVID-19 pandemic placed tremendous strain on everyone around the world. In order to prevent further transmission, strong preventive measures are intensified from the past weeks; however, the number of infected cases are consistently increasing around the world, even after undergoing lockdown.

Mathematical approaches are widely used to infer critical epidemiological transitions and parameters of COVID-19. Methods such as epidemic curve fitting, surveillance data during the early transmission R_0,_ and other epidemic models are frequently applied to generate forecasts of COVID-19 pandemic across the world [4-6].

## Methods

We used the data of cumulative confirmed death and recovery of COVID-19 cases reported from January 21 until March 26, 2020, that were obtained from John Hopkins Coronavirus resource center (https://coronavirus.jhu.edu/). We analyzed the data using dynamic models to generate 30 days forecasts and to understand the positive effect in the near future as well as projecting trends over trajectories. We used different statistical phenomenological models in the R-language platform to analyze the disease-based trajectories model for prediction purposes. We precisely used four models to analyze the aggregate data set for time series analysis. This includes the ARIMA model, which is a mass model of two different models, including the AR model (Auto Regression) and the MA (Moving Average) model [7]. This model results over Akaike information criterion (AIC) statistics and coverage of regression analysis.

Another type of COVID-19, like SARS disease (Severe Acute Respiratory Syndrome), is analyzed without breaking the current situation and predicting the future perspective [8]. VAR model (Vector Auto Average) model is used to predict the spatial extinct while using remote sensing data and for the purpose of creation of GIS map of worldwide on three different variables [9]. These three variables in the GIS environment create a map of cumulative confirmed cases country-wise as well as recovered and death map [10]. The use of another statistical analysis is a generalized logistic growth model (GLM), which generally depicted as a scaling parameter of integrating an additional result-oriented value put method [11]. Some epidemic models used in disease epidemic conditions measure oscillates some multiple peak parameters inferred in sub-epidemic and pandemic conditions to determine the projected outcomes [12].

After standardizing all the models, the data of the top 20 countries were included to analyze the forecasting models of differential spatial adjacent, and projected trajectories are analyzed up to April 30, 2020. By using GIS and Remote sensing to determine the epidemic mapping and analyzing the upcoming effects of COVID-19.

## Results and discussion

An overview of epidemic trends of COVID-19 across 170 countries and territories from January to March 26, 2020, is presented in Figure 1. From these top 20 countries, with a high number of confirmed cases are stratified to include in mathematical mode. The comparison between cumulative incident cases, mortality, and recovery of COVID-19 information among the top 15 affected countries is shown in Figure 2. Top countries’ data of China, Italy, Spain, and Iran showed highly disastrous mortality and badly effected with a vast number of COVID-19 cases. On the other hand, some European countries such as Switzerland, United Kingdom, Netherlands, Austria, Belgium, Norway, and Sweden had huge confirmed cases but negligible number in case of recover and death cases.

**Fig. 1:**
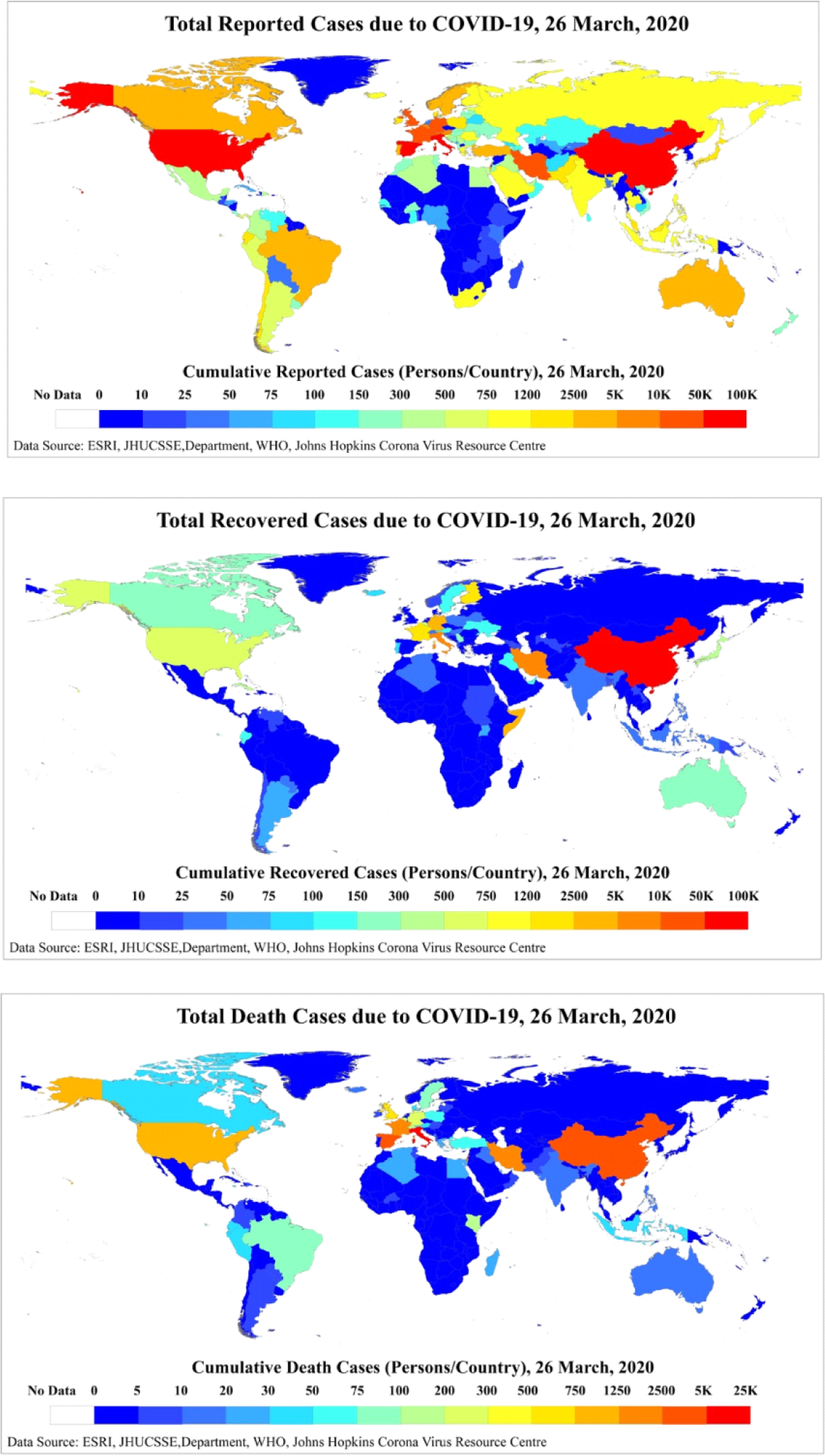
COVID-19 Epidemic Spatial pattern of total confirmed, recovery and death cases from 19 January to 26 March 2020 by Country, Territory, or Conveyance.

**Fig. 2:**
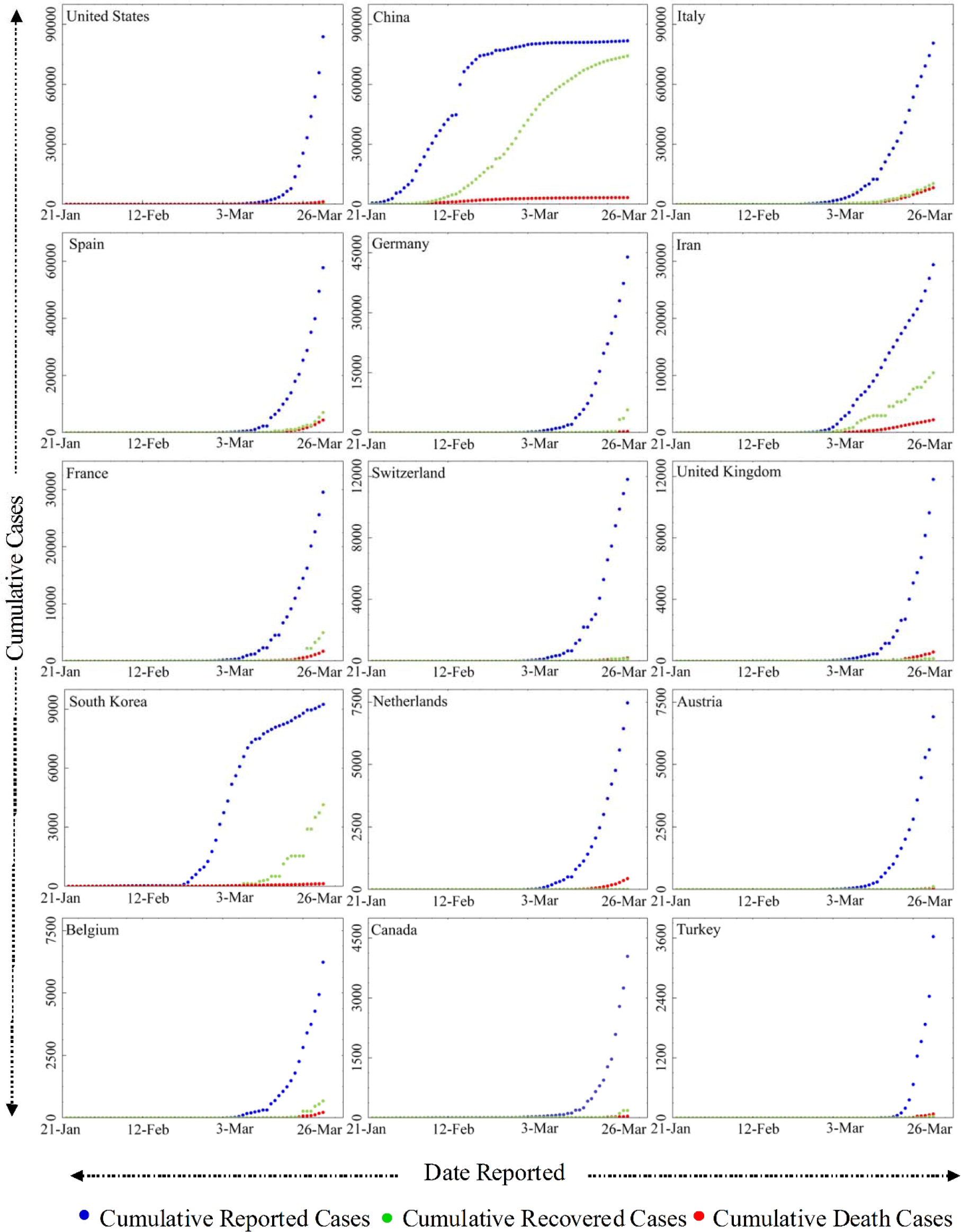
Comparisons between cumulative reported, recovery and death incidence of cases with COVID-19 on the top 15 affected countries.

Furthermore, data smoothening was applied to stabilize the data by removing changes in the level of a time series, and therefore eliminating (or reducing) trend and seasonality. After this, the forecast prediction model was applied by using AR and MA models to generate plots of the different trends in upcoming days. The outcome of these predictions is presented in Figure 3. Our findings revealed linearity in the current cumulative cases and showed a rapid exponential growth phase in the world span may occur roughly during April 8 to April 30, 2020, when the number of COVID-19 cases may skyrocket near to one million in the USA, 300,000 in Italy, and 250,000 in Germany, 150,000 in the United Kingdom and Iran (120,000). Other countries with a smaller number of cases but showing a sharp upward trend include Switzerland, Austria, and Canada. However, the cases of COVID-19 in China and South Korea remain stable. Public health officials in these countries need to grasp the powerful wave of exponential growth before COVID-19 collapses the entire health system.

**Fig. 3:**
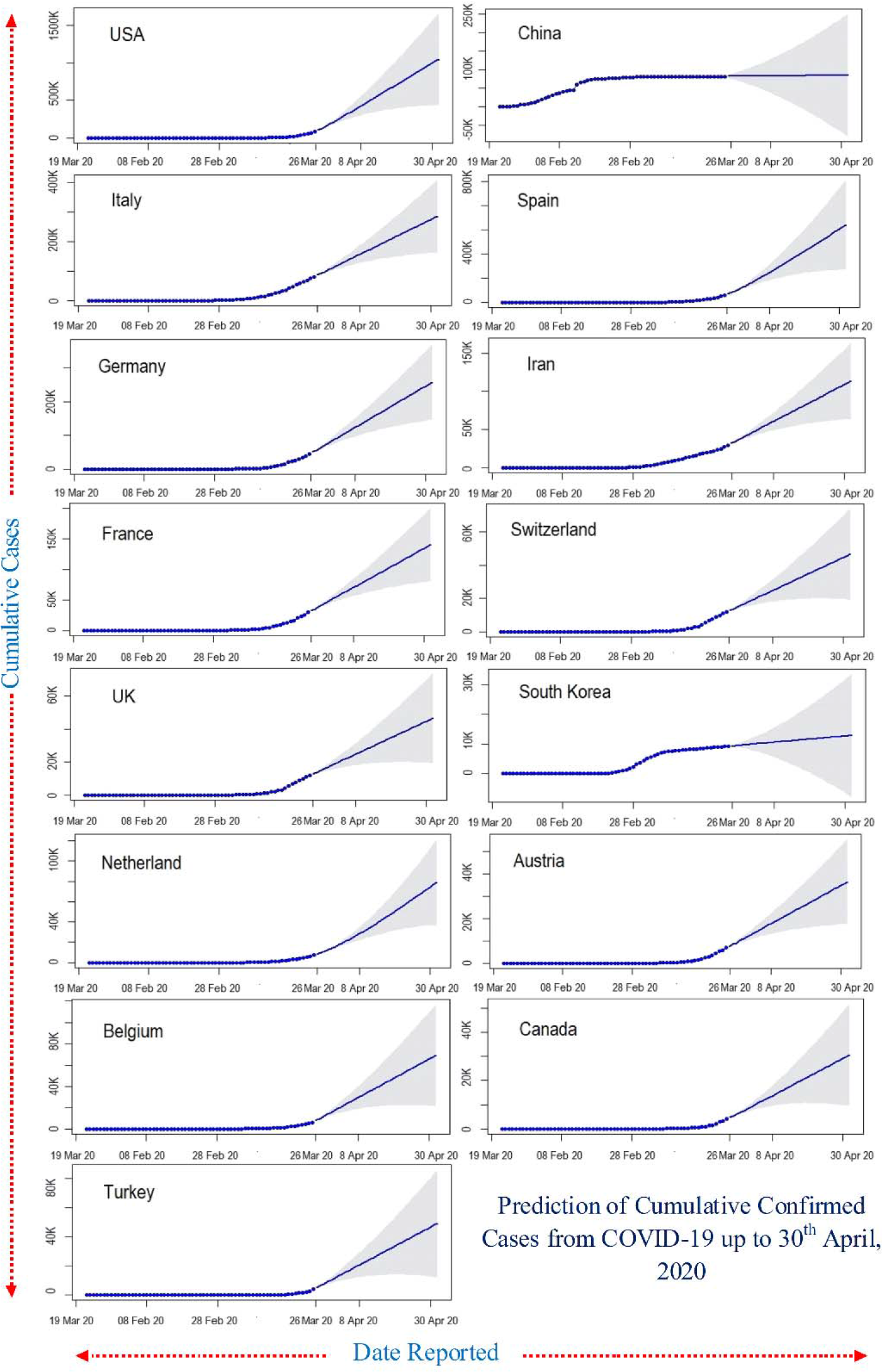
Twenty-day ahead ARIMA model forecasts of cumulative confirmed COVID-19 cases in the top 15 affected countries generated on 26 March, 2020. The light black line correspond to the cumulative cases confirmed up until 26 March 2020; the dark blue lines correspond to the mean ARIMA model; the light shadow lines depict the 95% prediction intervals and forecasting periods.

The fast raising COVID-19 infection rates may lead to the exponential death toll in the top 15 countries. Our analysis detected that the number of deaths would increase more rapidly in thousands during the mid of April 2020 (Figure 4). The associated mortality rate will be much higher in Italy, and Spain with roughly 30,000 deaths in each country, following France, USA and Iran will record around 10,000 deaths, and also the death in UK, Netherlands, Belgium, and Switzerland started to incline the death rates rapidly than the rest of current top 15 countries. The recovery rates will stay slow in initial time but then rapidly increase in Italy, Germany, Spain, and Iran by the end of prediction (Figure 5).

**Fig. 4:**
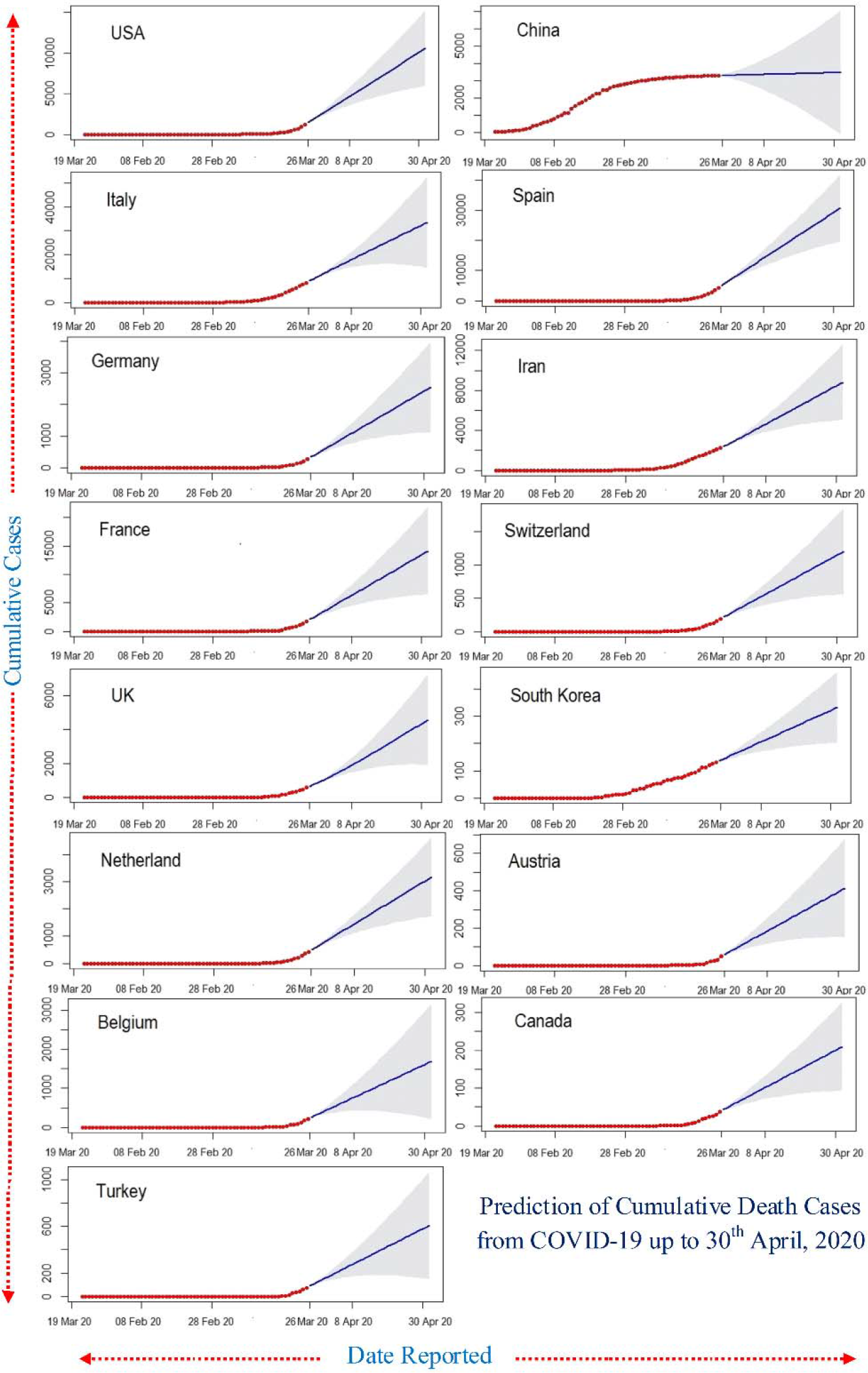
Twenty-day ahead ARIMA model forecasts of cumulative death COVID-19 cases in the top 15 affected countries generated on 26 March, 2020. The light black line correspond to the cumulative cases death up until 26 March 2020; the dark blue lines correspond to the mean ARIMA model; the light shadow lines depict the 95% prediction intervals and forecasting periods.

**Fig. 5:**
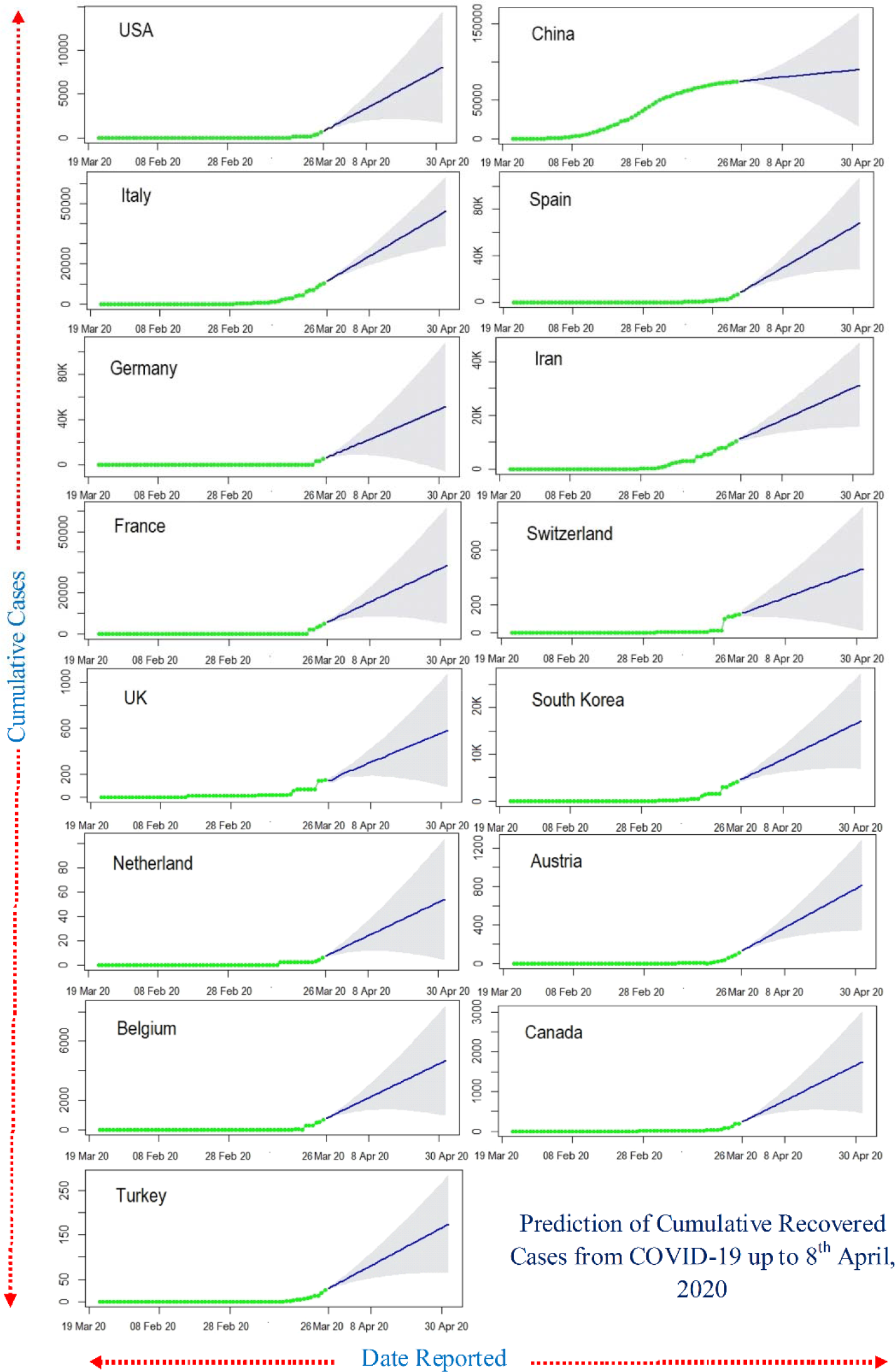
Twenty-day ahead ARIMA model forecasts of cumulative recovered COVID-19 cases in the top 15 affected countries generated on 26 March, 2020. The light black line correspond to the cumulative cases recovered up until 26 March 2020; the dark blue lines correspond to the mean ARIMA model; the light shadow lines depict the 95% prediction intervals and forecasting periods.

Our forecast analysis of COVID-19 dynamics showed a different angle for the whole world, and it looks scarier than imagined. Overall, our analysis confirmed the with an increase in the number of cases, the number of deaths is going to rise exponentially to rising in the future time (Figure 6). Interestingly, the recovery numbers also look promising, going to pack up during mid-April. Thus, it can slow down the surge of COVID-19 pandemic by the end of April.

**Fig. 6:**
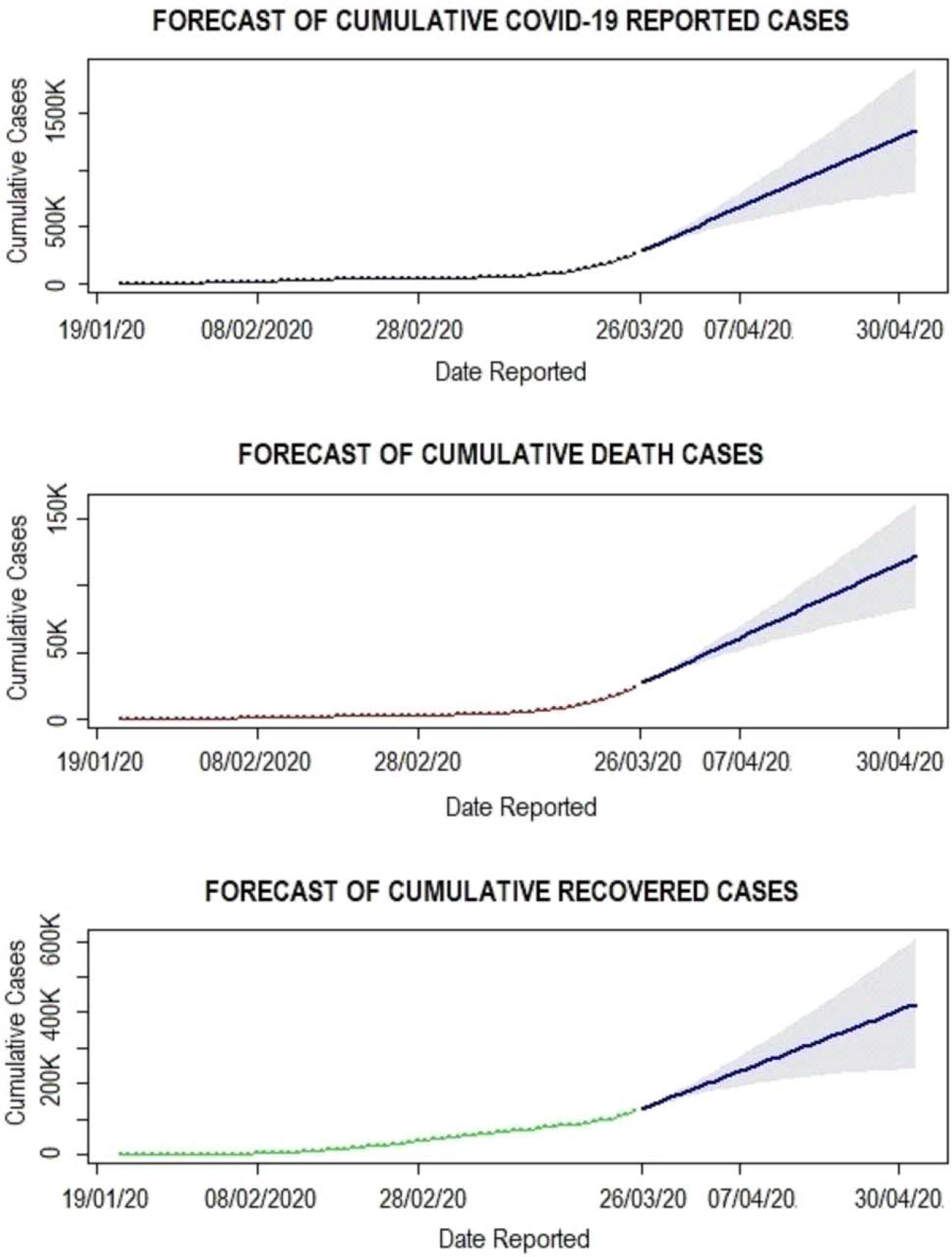
Twenty-day ahead ARIMA model forecasts of cumulative confirmed death and recovered Novel Coronavirus Disease (COVID-19) cases by Country, Territory, or Conveyance generated on 26 March, 2020. The red line correspond to the cumulative confirmed death and recovered up until 26 March 2020; the blue lines correspond to the mean ARIMA model; the light shadow lines depict the 95% prediction intervals and forecasting periods.

## Conclusion

Based on our predictions, public health officials should tailor aggressive interventions to grasp the power exponential growth, and rapid infection control measures at hospital levels are urgently needed to curtail the COVID-19 pandemic.

## Data Availability

All the data was obtained from the open-source repository

## Conflict of interest

None

## Notes

### Competing Interest Statement

The authors have declared no competing interest.

